# COVID-19 in Portugal: predictability of hospitalization, ICU and respiratory-assistance needs

**DOI:** 10.1101/2020.09.29.20203141

**Authors:** Andre Patricio, Rafael S. Costa, Rui Henriques

## Abstract

The current SARS-COV-2 epidemic is associated with nearly 1 million estimated deaths and responsible for multiple disturbances around the world, including the overload of health care systems. The timely prediction of the medical needs of infected individuals enables a better and quicker care provision for the necessary cases, supporting the management of available resources.

This work ascertains the predictability of medical needs (as hospitalization, respiratory support, and admission to intensive care units) and the survivability of individuals testing SARS-CoV-2 positive considering a cohort with all infected individuals in Portugal as per June 30, 2020. Predictions are performed at the various stages of a patient’s cycle, namely: pre-hospitalization (testing time), pos-hospitalization, and pos-intensive care. A thorough optimization of state-of-the-art predictors is undertaken to assess the ability to anticipate medical needs and infection outcomes using demographic and comorbidity variables, as well as onset date of symptoms, test and hospitalization.

## 1 Introduction

The novel coronavirus (COVID-19) is a disease caused by the Severe Acute Respiratory Syndrome Coronavirus 2 (SARS-CoV-2) infection, transmissible person-to-person and associated with acute respiratory complications in severe cases [5, 24]. The main symptoms of patients infected are fever, cough and tiredness, others are asymptomatic [10]. SARS-COV-2 pandemic presents an important threat to global health and is directly responsible for many deaths. Since the first outbreak (December 2019) in Wuhan, China, the number of confirmed patients infected worldwide has exceeded 30 million cases and nearly 1 million people have died from COVID-19^1^. Current literature has shown that most common of the infected patients with specific comorbidities/preconditions (e.g. hypertension, respiratory problems, diabetes) and old age are expected to develop a more severe response to the infection, and may consequently need longer hospitalizations and intensive care [18, 16, 22]. Strict social confinement efforts to decrease the COVID-19 R0 value (average number of individuals infected by each infected person) and guarantee the optimal use of equipment and beds at normal, continuous and intensive care units (ICU). However, although the public health responses aimed at delaying the spread of the infection, several countries such as United States, Brazil, Italy and India have been faced with severe healthcare crisis.

Without effective antiviral drugs and a vaccine, effective prognoses of COVID-19 disease are required. Statistical and computational models could assist clinical staff in triaging patients at high risk for respiratory failure to better guide the allocation of medical resources. Recently, several predictive models ranging from statistical and score-based systems to more recent machine learning models have been proposed in response to COVID-19. Guan et al. [11] proposed a Cox Regression Model to infer potential risk factors associated with series adverse outcomes in patients with COVID-19. Univariate and Multivariate Logistic Regression models have been used to determine risk factors associated with mortality [9]. Scoring systems have been proposed to predict COVID-19 patient mortality but are limited by small sample sizes [17, 12]. Other statistical approaches have also been emerging to aid prognostics [1, 8, 13]. Complementarily, machine learning (ML) methods offer the possibility of modeling more complex data relationships, generally yielding powerful capabilities to predict outcomes of infectious diseases in medical practice [3, 14]. To this end, classification and regression models have been proposed to risk stratification of patients and screen the spread of COVID-19 [20, 6, 2]. Despite the inherent potentialities of ongoing efforts, the exiting studies in the context of COVID-19 are limited by the size of available cohorts, generally neglect the predictability of medical needs (instead the focus is commonly placed on measurable disease factors, early detection of infection, and mortality risk prediction [23, 4, 21]), and no one comprehensively targets the Portuguese population up to date.

This study provides a structured view on the predictability of hospitalizations, ICU internments, respiratory assistance needs and survivability outcomes using a retrospective cohort encompassing all individuals having tested SARS-CoV-2 positive at Portugal as per June 30, 2020.

To this end, and considering demographic, co-morbidity and care-provision variables collected for the infected individuals, an assessment methodology is conducted whereby state-of-the-art predictive models are hyperparameterized and robustly evaluated in order to assess the upper bounds on the predictive performance for each one of the targeted variables. In addition, whenever applicable, this analysis is extended towards the various stages of a patient’s cycle: pre-hospitalization (testing time), after hospitalization, and after ICU internment. The gathered results pinpoint the relevance of the optimized predictors as good candidates to support care decisions for the Portuguese population.

The work is structured as follows. Section 2 presents major results and implications on the predictability of healthcare needs and outcomes for the Portuguese case. Section 3 details the undertaken validation methodology. Finally, Section 4 provides concluding remarks from the conducted study.

## 2 Results and discussion

Results on the predictability of the hospitalization needs (section 2.2), ICU internments (section 2.3), respiratory assistance (section 2.4) and outcome (section 2.5) of infected individuals at Portugal – as of June 30, 2020 – are gathered and discussed below.

### Data source

A retrospective cohort (from February to June 30, 2020) of confirmed COVID-19 patients in Portugal was used for this study. The anonymized dataset was provided from Direção-Geral da Saúde (DGS). The gathered data, termed covid19-DGS database, contains information pertaining to the demographic, clinical patient characteristics and preexisting conditions.

### Ethical considerations

The COVID-19 dataset was provided by DGS under a collaboration in the context of the score4COVID research project. The conducted tasks along the score4COVID project were further validated by the Ethical Committee of the FCT-NOVA University.

### 2.1 Cohort study

The target cohort consists of a total of 38.545 individuals with SARS-CoV-2 positive, which encompasses until June 30, 2020: 17.046 recoveries (SARS-CoV-2 negative after positive testing) and 1.155 deaths. Within the target population, we find 4.327 hospitalizations, and 253 internments in the ICU. Among ICU internments, thee are 82 recoveries and 61 deaths. Regarding the needs for respiratory support, we find a total of 180 individuals that undertook assisted ventilation, 292 individuals submitted to oxygen therapy, and 9 individuals with alternative modes of respiratory support (e.g. extracorporeal membrane oxygenation).

The major classes of comorbidities monitored are neoplasm, diabetes, asthma, pulmonary, hepatic, hematological, renal, neurological, neuromuscular and immunodeficiency conditions. The representatitivity of individuals with one or more comorbidities, as well as their impact on survivability, are depicted in Figure 2. Figure 1 further provides additional statistics, including the sex and age distributions for the target population, and the average number of days from onset symptoms (traced by the public health line for COVID-19) to positive testing and hospitalization.

**Figure 1:**
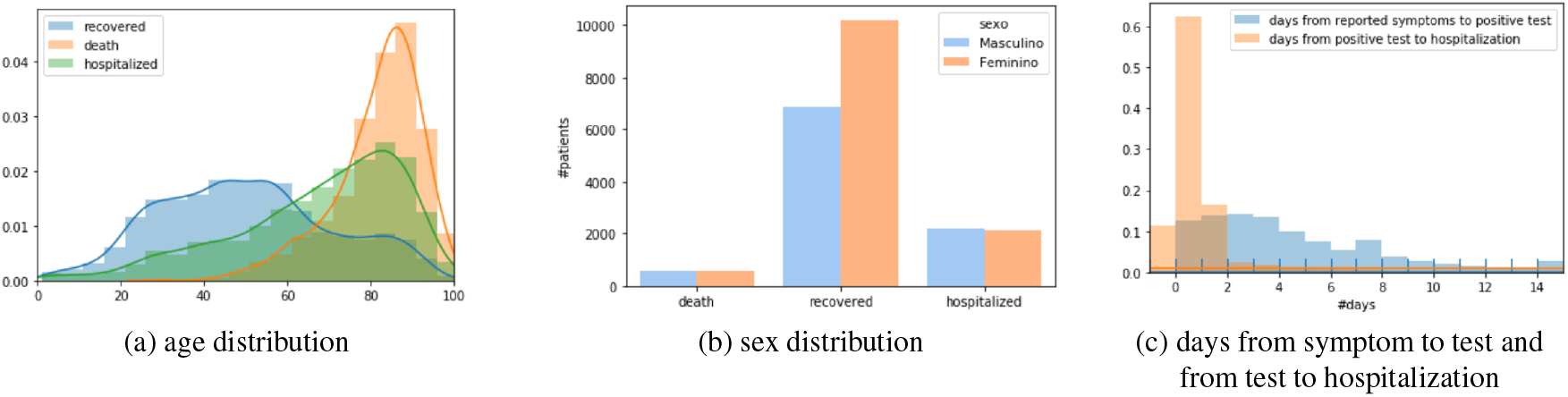
Cohort statistics: a-b) demographic distribution of infected individuals with known outcome – death and recovery – and hospitalized (containing both cases with and without a known outcome); c) average number of days between care stages (plotted negative bin corresponds to hospitalizations before SARS-CoV-2 testing).

**Figure 2:**
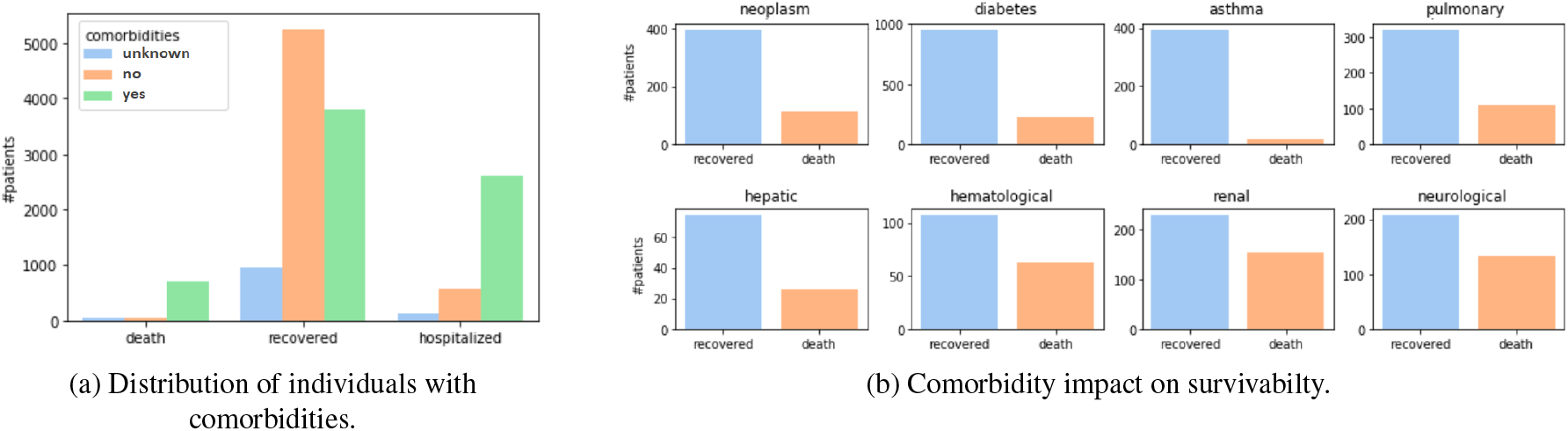
Cohort statistics: a) distribution of individuals with one or more comorbidities among deaths, recovered cases and hospitalizations; b) association between individual comorbidities and survivability outcomes..

### 2.2 Hospitalization

Figure 3 provides results on the ability to predict the need for an individual to be hospitalized once tested as SARS-CoV-2 positive, when considering the: i) demographic group (age and gender), ii) co-morbidity fac-tors. Co-morbidity factors are categorized in accordance with the presence/absence of kidney, asthma, lung, cancer, neuromuscular, diabetes, HIV, cardiac, and pregnancy conditions. Understandably, individuals without SARS-CoV-2 negative testing after infection were excluded from this analysis. The data preprocessing, classifier hyperparameterization and validation methodology are detailed in section 3.

**Figure 3:**
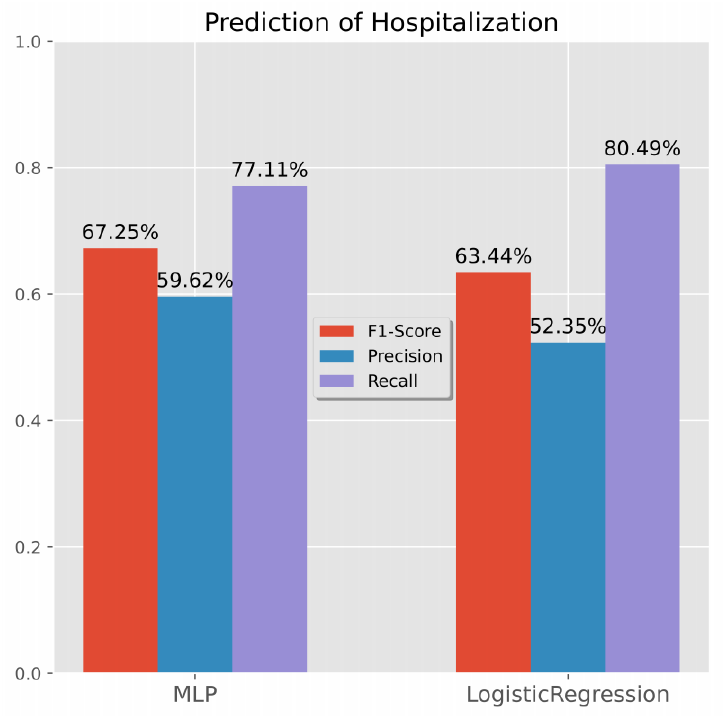
Predictability of hospitalizations for individuals testing SARS-CoV-2 positive. Recall, precision and F1 for the best predictors in F1 (*left*) and recall (*right*) scores on validation set after 10CV hyperparameterization.

Generally, we observe that over 75% of the hospitalization needs can be identified at the SARS-CoV-2 testing time. This level of recall/sensitivity is observed at the expense of a precision around 50%, meaning that half of the predicted hospitalization needs may not be observed in practice. Multilayer percepton networks and logistic regressors were the best performing classification models according to F1-score and recall, respectively. These results provide empirical evidence towards the role of these predictors in supporting monitoring decisions.

### 2.3 ICU internment

Figure 4 assesses the ability to anticipate intensive care needs for infected individuals at two times: a) before hospitalization, and b) after hospitalization. To this end, section 3 methodology was pursued considering demographic factors, co-morbidity factors, and the time-to-hospitalization for hospitalized individuals. Individuals without SARS-CoV-2 negative testing after infection are excluded.

**Figure 4:**
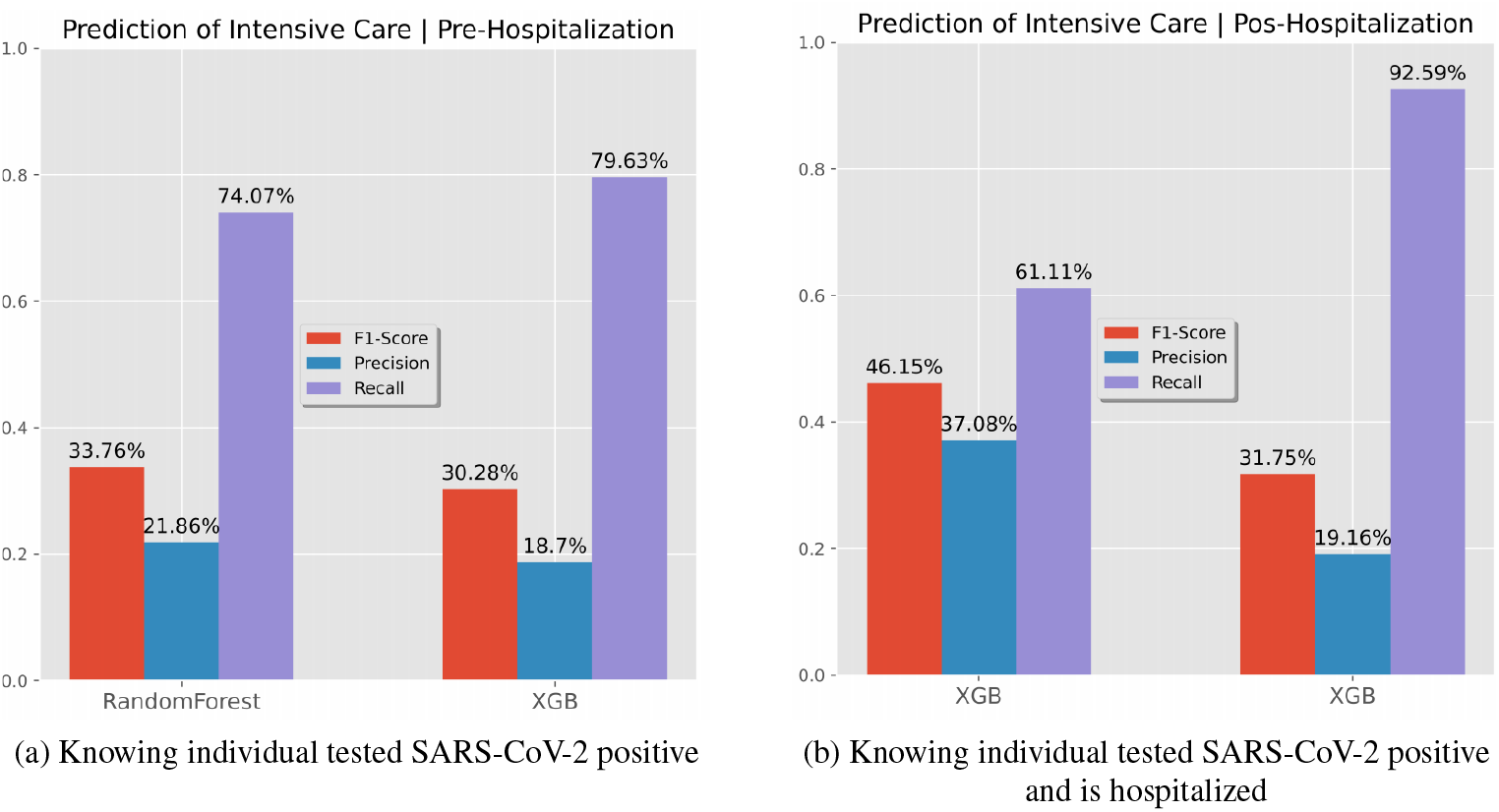
Predictability of ICU internment. Results for the best F1 predictor (*left*) and recall predictor (*right*).

The predictability of ICU needs are less satisfactory for both pre- and pos-hospitalization testing scenarios. We hypothesize that the difficulty of predicting ICU needs is partially related with the small number of individuals with ICU internments in our dataset, together with the presence of missing values associated with ICU internment needs for most individuals. Even though we can achieve a recall of 92.6% with Gradient Boosting (XGBoost) in a Pos-Hospitalization setting, it comes with the cost of a quite low precision. Still, this classification model is suggested to support monitoring decisions at the hospital bedside, as its specificity is still considerably high.

### 2.4 Respiratory support

Figure 5 assesses respiratory assistance needs for hospitalized individuals with SARS-CoV-2, considering three assistance modes: i) ventilation support, ii) oxygen therapy, and iii) combined ventilation and oxygen therapies. To this end, section 3 methodology was pursued considering demographic, co-morbidity and time-to-hospitalization factors. Individuals without SARS-CoV-2 negative testing after infection were excluded from this analysis. As respiratory support is a multiclass variable, we now consider different performance evaluation by focusing on the: i) recall for each major class (ventilation, oxygen and non-required support), and ii) precision of individuals with oxygen or ventilation assistance.

**Figure 5:**
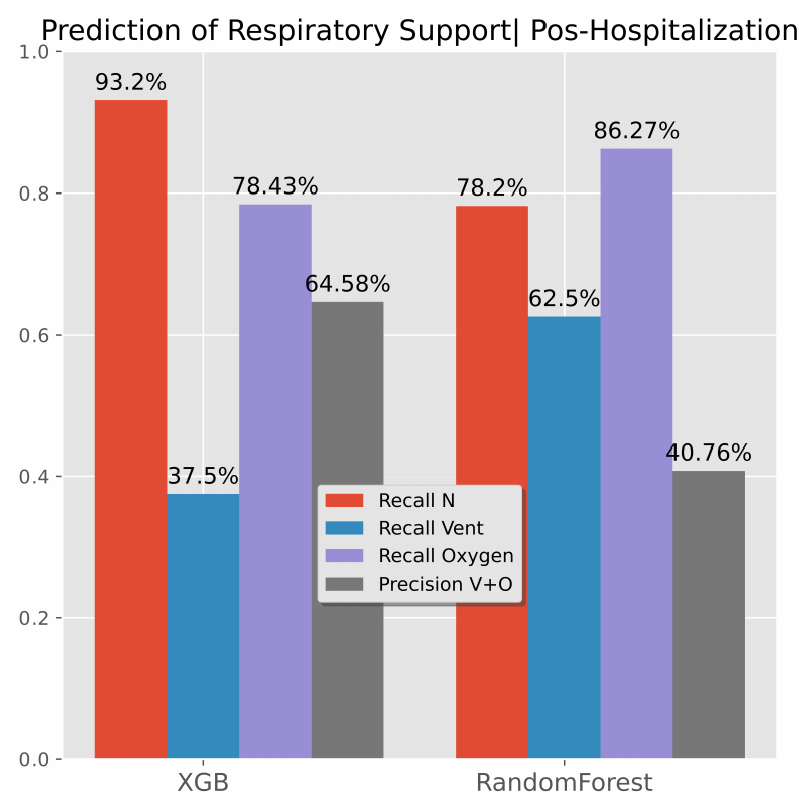
Predictability of respiratory support needs – assisted ventilation, oxygen therapy and combined support – for hospitalized individuals with SARS-CoV-2. Performance of the best F1 predictor (*left*) and recall predictor (*right*).

RandomForests can attain a satisfactory identification of hospitalized individuals that may require respiratory support in the future, generally providing recalls for each assistance mode above 60% at the cost of a 40% precision. According to the conducted methodology, they are pinpointed as a good candidate to support in-hospital care decisions.

### 2.5 Survivability (outcome)

Finally, Figure 6 provides an analysis of the ability to predict the outcome (recovered versus death) for individuals with SARS-CoV-2 infection at three different times: i) before hospitalization (testing time), ii) after hospitalization, and iii) after ICU internment when applicable. To this end, we preserved the input variables and validation methodology (section 3) considered in previous scenarios.

**Figure 6:**
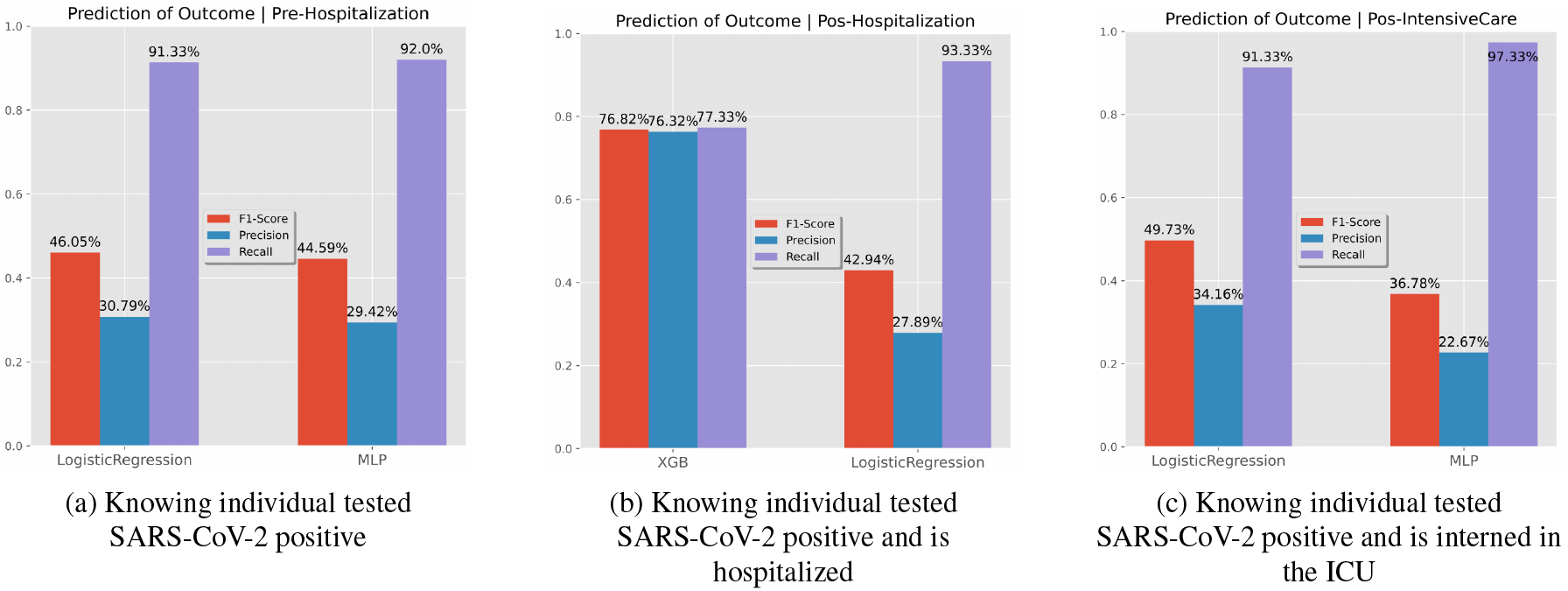
Predictability of the survivability (outcome) of infected individuals before hospitalization, after hospitalization and after ICU interment. Results for the best F1 predictor (*left*) and recall predictor (*right*).

Our results show a high ability to identify death outcomes, although at a cost of wrongly classifying two thirds of individuals susceptible to death. In the pos-Hospitalization scenario we achieve more balanced results, with both precision and recall around 75% using Gradient Boosting. The introduction of the intensive care variable ends up hampering results since it offers both a view on acute needs as well as continuous care instruments.

## 3 Methods

Complete subpopulations from the target cohort are produced for each target variable by guaranteeing the presence of all individuals undertaking the target form of care (hospitalization, ICU internment, respiratory support) as well as all remaining individuals with recovery-or-death outcomes. After this data curation step, we proceed with the optimization of data preprocessing options and classifiers’ selection hyperparameters for each of the target variables separately.

To this end, we applied a nested 10-fold cross-validation methodology, whereby we first create train-test partitions (outer cross-validation) to assess the performance of an optimized classification method, and within each training fold we further create train-test partitions (inner cross-validation) for hyperparameterizing the classifier under assessment. This methodology guarantees that all observations are used to assess the final performance and prevents biases as hyperparameterization takes place within each training folds.

Within each inner train-test fold, Bayesian Optimization (BO) [19] is applied to find the hyperparameters that best fit the pipeline. The optimization measures are:

– F1-Score and recall for binary classes; and
– Cohen-Kappa and average class recall for target variables with more than 2 classes (respiratory support).

The selected views imply that each target variable has two different sets of optimized classifiers (as shown in Figures 3 to 6): one that aims to weight recall-precision views and other optimized towards the true positive rate irrespective of the positive predictive value (precision).

The allowed preprocessing options are: imputation of missing values using median-mode imputation, KNNImputer or none; class balancing using subsampling, oversampling, SMOTE or none; and normalization of real-valued variables using standardization, scaling or none. The selected classifiers are: Bernoulli Naive Bayes, Gaussian Naive Bayes, K-Nearest Neighbors, Decision Tree, Random Forest, XGBoosting, Logistic Regression, Multi-Layer Perceptron. We considered the implementations provided in scikit-learn [15] and xgboost [7] packages in python.

For each classifier, all the supported parameters in scikit-learn were subjected to hyperparameterization. Regarding the Multi-Layer Perceptron, we placed upper limits on the number of hidden layers (three) and nodes per layer (twenty) given the low-dimensionality nature of the target dataset. The hyperparameters were subjected to a total of 50 iterations.

## 4 Concluding remarks

This work offers a discussion on the predictability of hospitalization needs, ICU internments, respiratory assistance and survivability outcome of individuals infected with SARS-CoV-2 at Portugal as of June 30, 2020. A retrospective cohort with all confirm COVID-19 cases since February, encompassing demographic and co-morbidity variables, is considered as the target population to this end. This study has some inherent limitations: i) the number of variables for the outcomes of interest were limited (e.g. the body mass index is missing) and ii) further external validation is required. However, we can anticipate that as more data are made available, we will be able to improve the performance of the models.

The gathered results for the given cohort study reveal that: 1) over 75% of hospitalization needs can be identified at the SARS-CoV-2 testing time (with *>*50% precision); 2) ICU needs are generally less predictable at both pre- and pos-hospitalization stages under the given cohort; 3) respiratory assistance needs (including ventilation support, oxygen therapy, and combined ventilation-oxygen support) achieve recall levels above 60% (with *>*40% precision); 4) death risk along different stages (testing time, after hospitalization and after ICU internment) has the highest degree of predictability.

The predictive models yielding better accuracy performance were associative classifiers, particularly XG-Boost and RandomForests, neural networks with hyperparameterized architectures and logistic regressors; with the optimal choice varying in accordance with the target variable and evaluation measure.

As future work, and in collaboration with DGS, our predictors are expected to be integrated within a clinical decision support system to be provide to the National Health Service (SNS).

The conducted study pinpoint the relevance of the proposed predictive models as good candidates to support medical decisions for the Portuguese population, including both monitoring and in-hospital care decisions. Knowing the most probable outcomes along the lifecycle of an SARS-CoV-2 infected individual can guide medical teams to act quickly near more vulnerable patients.

## Data Availability

The COVID-19 dataset was provided by DGS.

## Acknowledgments

The authors thank Direção Geral da Saúde (DGS) for providing data. This work was further supported by Fundação para a Ciência e a Tecnologia (FCT) under the LAETA (UIDB/50021/2020), IPOscore (DSAIPA/DS/0042/2018), Data2Help (DSAIPA/AI/0044/2018) and iCare4U (PTDC/EME-SIS/31474/2017) projects, the contract CEECIND/01399/2017 and by FCT/MCTES funds for the Associate Laboratory for Green Chemistry LAQV (UIDB/50006/2020).

## Footnotes

1 https://www.who.int (accessed September, 2020)

